# The COVID-19 Pandemic Vulnerability Index (PVI) Dashboard: Monitoring county-level vulnerability using visualization, statistical modeling, and machine learning

**DOI:** 10.1101/2020.08.10.20169649

**Authors:** Skylar W. Marvel, John S. House, Matthew Wheeler, Kuncheng Song, Yihui Zhou, Fred A. Wright, Weihsueh A. Chiu, Ivan Rusyn, Alison Motsinger-Reif, David M. Reif

## Abstract

**Background:** While the COVID-19 pandemic presents a global challenge, the U.S. response places substantial responsibility for both decision-making and communication on local health authorities, necessitating tools to support decision-making at the community level.

**Objectives:** We created a Pandemic Vulnerability Index (PVI) to support counties and municipalities by integrating baseline data on relevant community vulnerabilities with dynamic data on local infection rates and interventions. The PVI visually synthesizes county-level vulnerability indicators, enabling their comparison in regional, state, and nationwide contexts.

**Methods:** We describe the data streams used and how these are combined to calculate the PVI, detail the supporting epidemiological modeling and machine-learning forecasts, and outline the deployment of an interactive web Dashboard. Finally, we describe the practical application of the PVI for real-world decision-making.

**Results:** Considering an outlook horizon from 1 to 28 days, the overall PVI scores are significantly associated with key vulnerability-related outcome metrics of cumulative deaths, population adjusted cumulative deaths, and the proportion of deaths from cases. The modeling results indicate the most significant predictors of case counts are population size, proportion of black residents, and mean PM_2.5_. The machine learning forecast results were strongly predictive of observed cases and deaths up to 14 days ahead. The modeling reinforces an integrated concept of vulnerability that accounts for both dynamic and static data streams and highlights the drivers of inequities in COVID-19 cases and deaths. These results also indicate that local areas with a highly ranked PVI should take near-term action to mitigate vulnerability.

**Discussion:** The COVID-19 PVI Dashboard monitors multiple data streams to communicate county-level trends and vulnerabilities and facilitates decision-making and communication among government officials, scientists, community leaders, and the public to enable effective and coordinated action to combat the pandemic.

## Introduction

Defeating the COVID-19 pandemic requires well-informed, data-driven decisions at all levels of government, from federal and state agencies to county health departments. Numerous datasets are being collected in response to the pandemic, enabling the development of predictive models and interactive monitoring applications (Wynants et al. 2020; ESRI 2020). However, this multitude of data streams—from disease incidence to personal mobility to comorbidities—is overwhelming to navigate, difficult to integrate, and challenging to communicate. Synthesizing these disparate data is crucial for decision-makers, particularly at the state and local levels, to prioritize resources efficiently, identify and address key vulnerabilities, and evaluate and implement effective interventions. To address this situation, we developed a COVID-19 Pandemic Vulnerability Index (PVI) Dashboard (https://covid19pvi.niehs.nih.gov/) for interactive monitoring that features a county-level Scorecard to visualize key vulnerability drivers, historical trend data, and quantitative predictions to support decision-making at the local level (Figure 1).

**Figure 1.**
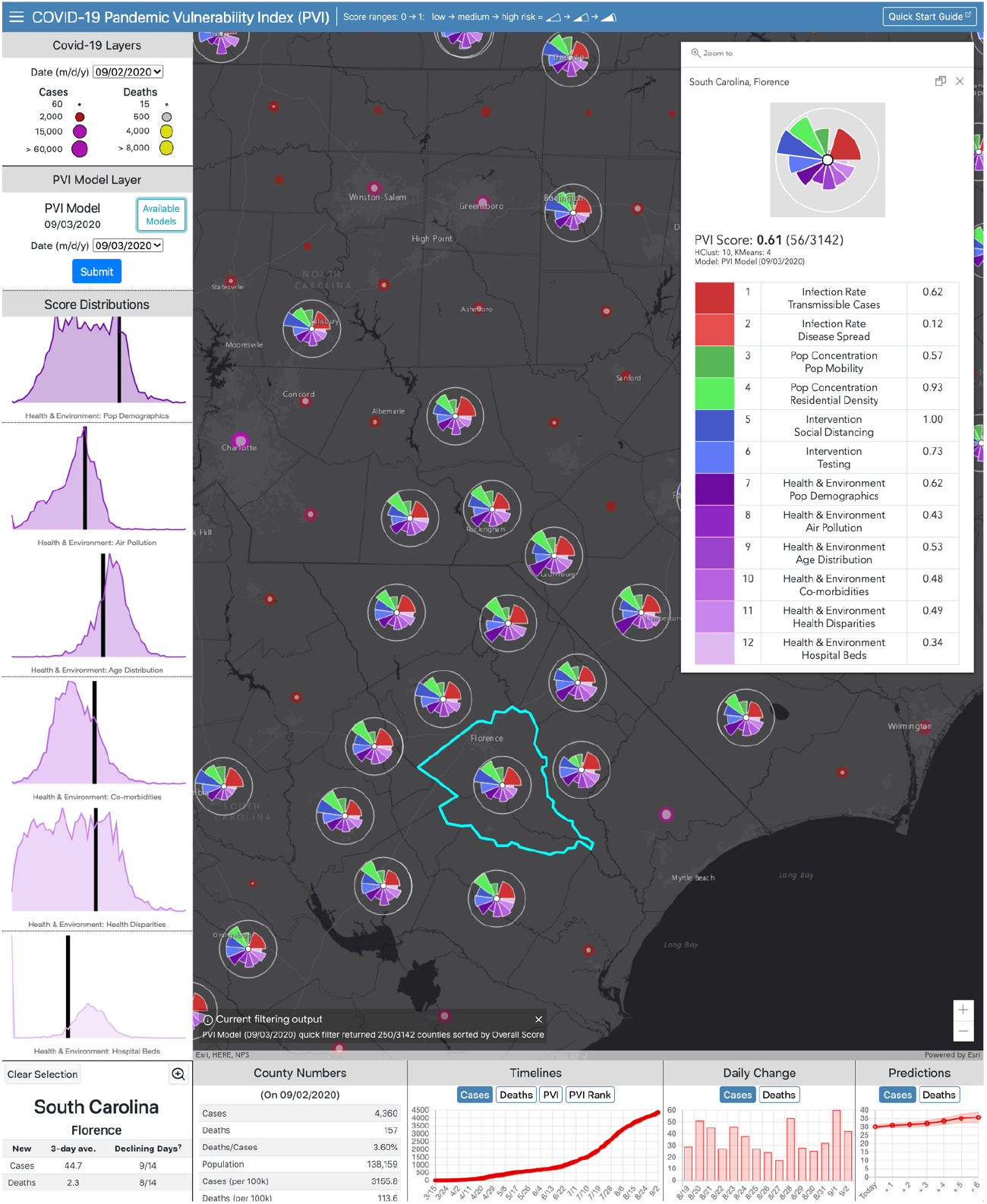
Dashboard displaying map view with PVI Scorecard and associated data for a selected county. The Dashboard allows U.S.-wide navigation to area(s) of interest. The filter is set to display the top 250-ranked (i.e., most vulnerable) PVI profiles for the selected date (displayed in the upper left panels for each data layer). The Scorecard displayed shows the contribution of each indicator (slice) for Florence, SC, which is in a cluster of high-PVI counties across the rural Southeast U.S. The scorecard summarizes the overall PVI score and rank compared to all 3,142 U.S. counties. In the graphical profile, longer slices indicate higher vulnerability driven by a particular indicator, with corresponding indicator-wise scores (0 = minimum; 1 = maximum) provided in the lower portion of the Scorecard. The scrollable score distributions at left compare the selected county PVI to the distributions of overall and slice-wise scores across the U.S. The panels below the map are populated with county-specific information on observed trends in cases and deaths, observed numbers for the selected date, historical timelines (for cumulative cases, cumulative deaths, PVI, and PVI rank), daily case and death counts for the most recent 14-day period, and a 7-day forecast of predicted cases and deaths. The information displayed for both observed COVID-19 data and PVI layers is scrollable back through March 2020. Documentation of additional features and usage, including advanced options (accessible via the collapsed menu at the upper left), is provided in a Quick Start Guide (linked at the upper right corner).

We assembled U.S. county- and state-level datasets into 12 key indicators across four major domains: current infection rates (infection prevalence, rate of increase), baseline population concentration (daytime density/traffic, residential density), current interventions (social distancing, testing rates), and health and environmental vulnerabilities (susceptible populations, air pollution, age distribution, comorbidities, health disparities, and hospital beds). These 12 indicators (some of which combine multiple datasets) are integrated at the county level into an overall PVI score, employing methods previously used for geospatial prioritization and profiling (Bhandari et al. 2020; Marvel et al 2018). The individual data streams comprising these indicators measure either well-established, general vulnerability factors for public health disasters or emerging factors relevant to the COVID-19 pandemic (Centers for Disease Control and Prevention 2015).

In developing the PVI, we performed rigorous statistical modeling of the underlying data to enable quantitative analysis and monitoring and provide short-term predictions of cases and deaths. Our modeling efforts directly address the discussion raised by Chowkwanyun and Reed (2020) about racial disparities in COVID-19 case and death rates. By contextualizing factors such as these racial disparities, correcting for socioeconomic factors, health resource allocation, and co-morbidities, and highlighting place-based risks and resource deficits, the PVI can help explain differences in the spatial distribution of cases. Specifically, we performed three types of modeling efforts, all of which are regularly updated. First, epidemiological modeling on cumulative case- and death-related outcomes provides insights into the epidemiology of the pandemic. Second, dynamic time-dependent modeling provides similar outcome estimates as national-level models but with county-level resolution. Finally, a Bayesian machine learning approach provides data-driven, short-term forecasts. Herein, we describe the development of the PVI, including the epidemiological modeling and machine-learning forecasts, and its use in an interactive web Dashboard.

## Methods

### Data Streams Included in the Pandemic Vulnerability Index

To the best of our knowledge, we have assembled the most extensive set of community-level data streams related to COVID-19. These data streams span four major domains, namely infection rate, population concentration, intervention measures, and healthcare vulnerability. The specific components (i.e., datasets) comprising the current PVI model are provided in a dedicated Details page linked from the Dashboard. Supplementary Table 1 describes each component, outlines the rationale for its inclusion, and provides a link to the associated data source. To empower additional modeling efforts, the complete time series of all daily PVI scores and the source data are publicly available at https://github.com/COVID19PVI/data. The software used to generate PVI scores and profiles from these data is freely available at https://toxpi.org (Marvel et al. 2018).

These data streams comprise both static and dynamic data, including static measures of population concentration and healthcare vulnerabilities. Many of the data streams are from the CDC’s Social Vulnerability Index (SVI), which was developed by the Agency for Toxic Substances and Disease Registry (ATSDR’s) Geospatial Research, Analysis and Services

Program (GRASP). GRASP creates and maintains databases that help emergency response planners and public health officials identify and map the communities most likely to need support before, during, and after a disaster or hazard event such as the current pandemic. The SVI has been successfully used in a variety of emergency response scenarios, including mapping fire outbreaks to determine vulnerability metrics (Lue & Wilson 2017) and hazard mitigation planning studies (Horney et al. 2017). The CDC’s SVI uses U.S. Census data to rank each census tract’s social vulnerability based on 15 factors, including poverty, vehicle access, and housing crowding. Additional data streams from the 2020 County Health Rankings that summarize the prevalence of important co-morbidities and risk factors at the county level are also included (Bhandari et al. 2020). Regarding interventions, it has been established that increased testing rates and the implementation of social distancing are effective interventions to slow the spread of COVID-19 and fatalities due to the disease (Wu et al. 2020). Testing rates are from the COVID Tracking Project (The COVID Tracking Project 2020), and daily ordinal grades of social distancing adherence are from Unacast (2020), which analyzes relative mobility compared to the same period during the previous year using mobile device data. By anchoring movement to pre-COVID-19 activity, this measure is applicable in both rural and urban settings, which have different mobility patterns in general. Dynamic measures of disease spread and the number of transmissible cases are estimated from John Hopkins University data (Dong, Du, & Gardner 2020).

### PVI Calculation

For each county, the PVI, a dimensionless index score, is calculated as a weighted combination of all data sources and represents a formalized, rational integration of information from various domains. The score is calculated using the Toxicological Prioritization Index (ToxPi) framework for data integration, as described in Reif et al. (2010). Briefly, the individual factors are ranked for each county by scaling the raw value from 0 to 1. Factors for which lower values represent higher risk (percent testing and social distancing) are reverse-scaled so that higher values represent higher vulnerability. This allows all factors to be expressed on the same scale (0 to 1) and removes the difficulties associated with applying different units to different factors. The overall PVI is then calculated using a weighted sum. The choice of factors used in the creation of the PVI score and the weights applied to them were informed by our epidemiological modeling (described in subsequent section) as well as general knowledge of contributors to general health morbidities.

The PVI profiles translate numerical results into visual representations as component slices of a radar chart, with each slice representing one piece (or related pieces) of information. For each profile, the radial length of a slice represents its rank relative to all other entities (i.e., counties), with a longer radius indicating higher concern or risk. The relative width (e.g., fraction of a full, 360° circle) of a slice indicates the contribution of its score to the overall model. These visual profiles provide a risk assessment of the strength, relative contribution, and robustness of the multiple data sources used in the model. Figure 2 illustrates the PVI workflow and the results for two example counties. This type of data integration framework has been proven effective for communicating risk prioritization and profiling information among scientists, regulators, stakeholders, and the general public and has been featured in publications by the U.S. National Academy of Sciences, Engineering, and Medicine (2017) and the World Health Organization’s International Agency for Research on Cancer (Loomis et al. 2018).

**Figure 2.**
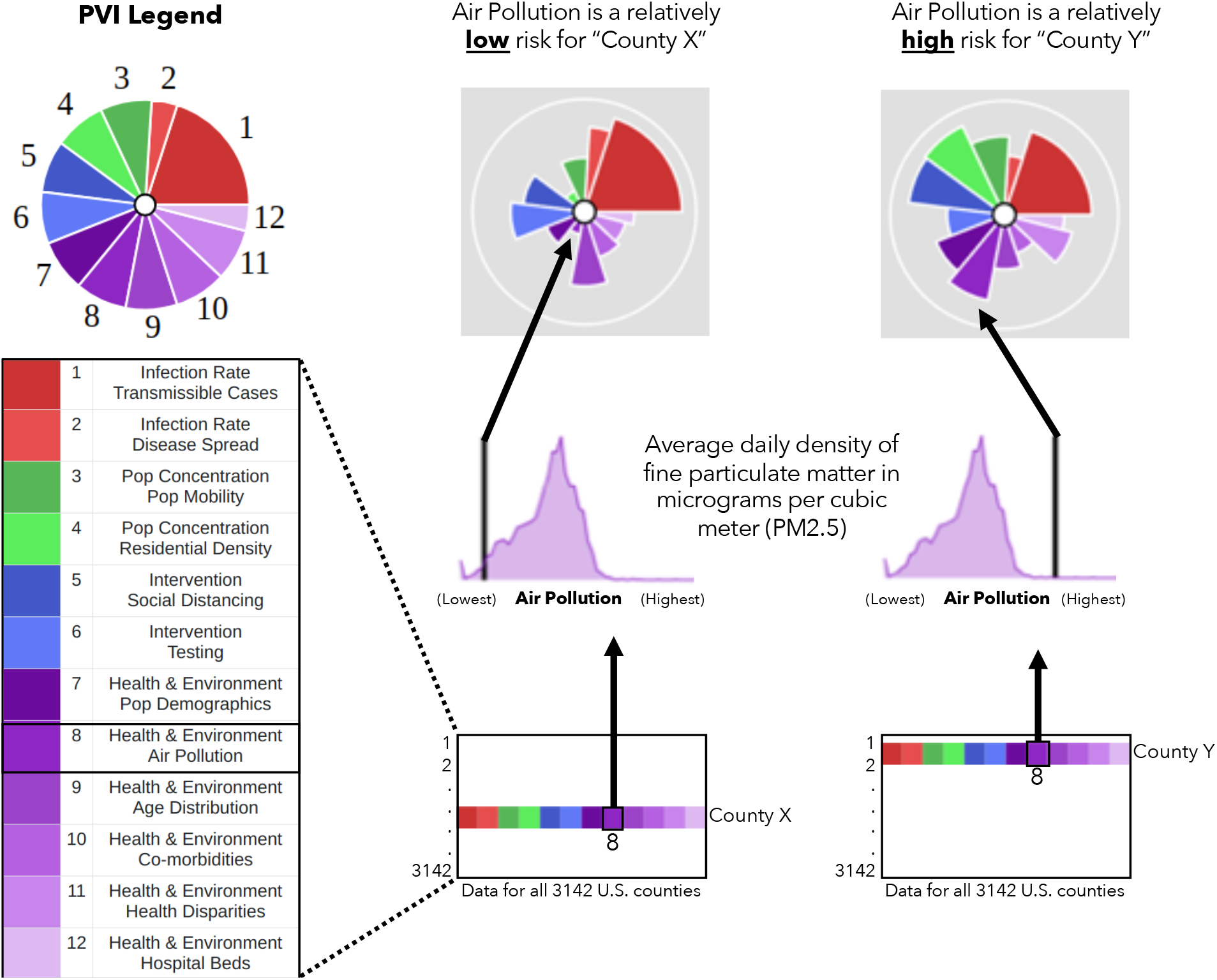
Translation of data into PVI profiles. Information from all 3,142 U.S. counties is translated into PVI slices. The illustration shows how *Air Pollution* data (average density of PM_2.5_ per county) are compared for two example counties. The county (County Y) with the higher relative measurement has a longer *Air Pollution* slice than the county (County X) with a lower measurement. This procedure is repeated for all slices, resulting in an integrated, overall PVI profile.

### Epidemiological Modeling

The diverse array of data assembled for the epidemiological modeling that informs the COVID-19 PVI Dashboard represents an advance over the ever-increasing number of models related to COVID-19. To provide context and ensure that the data streams provide conclusions and priority rankings that are broadly consistent with other epidemiological models, we performed cross-sectional analysis of cumulative (i) cases, (ii) deaths, (iii) deaths as a proportion of the population, and (iv) deaths as a proportion of reported cases using data current as of 8/24/2020. We emphasize that the PVI is not intended to be an epidemiological modeling tool per se as it does not explicitly distinguish between factors of vulnerability for cases vs. deaths. Our modeling described here is intended to anchor the components of the PVI and provide context within the larger field of COVID-19-related epidemiological modeling. Additionally, this modeling is not intended to provide forecasts, which are the primary focus of projection models, as discussed in the subsequent section (see Forecasting).

As the initial analyses displayed evidence of count overdispersion, we performed generalized linear modeling in R version 3.5 with the gam() procedure using a negative binomial model with observed cumulative counts as the response (see Supplementary Tables 2-5) (R Core Team 2018). For analyses (i), (ii), and (iv), we used log(population size) values as predictors with estimated coefficients. For analysis (iii), we used the “offset” command to model the death rate. Similarly, for analysis (iv), we used log(cumulative cases) as an offset to model the death rate among cases, which may produce biased results due to regional variation in reporting rates. It should be noted that a constant underreporting bias across counties would be absorbed into the intercept and would otherwise produce valid coefficient estimates for the predictors. Analysis (iv) may provide important clues about the death risk as including cases in the denominator removes a large portion of the stochastic variation. Moreover, for all analyses, we used the proportion of the state population that has been tested as a predictor to account for additional sources of bias.

To anchor our efforts to previous work, we included as additional fixed predictors those from Wu et al. (2020), who focused primarily on the effects of a PM_2.5_ air pollution index using an analysis analogous to our model (iii). Before analysis, we removed predictors with pairwise correlation with any other predictor greater than 0.85 and predictors that would be collinear with a series of predictors, such as the overall proportion of minority residents. For pairs exceeding the correlation threshold, we favored predictors with the lower missingness rate (if any) or those that are reported in other work. Dynamic predictors (i.e., those that changed substantially over the modeled period) were incorporated using simple county averages over the March-August period covered by the PVI. With over 3,100 counties (according to FIPS codes), most with >0 cases and deaths, the analysis can easily support the 27 to 28 final predictors used. To facilitate comparison with previous sources, we used predictors as they are given in their source. Accordingly, in some instances, predictors are represented as proportions and, in other instances, they are represented as percentages.

To provide additional context, we also performed negative binomial modeling (R version 3.5 bam() with “REML” fitting) (R Core Team 2018) of daily cases up to 6/11/2020 (Supplementary Table 6), using the fixed county predictors as well as unaveraged dynamic predictors. Due to the nature of the model, we included the two-week-lagged cumulative number of cases as an additional predictor, as well as a smoothing spline time-dependent term to reflect a nationwide component of risk. Although it is formally a fixed-effects model, we refer to this model as dynamic and treat each day outcome as an independent realization, with the rate determined by the predictors. To account for potential time-dependent latent correlation structures, we determined standard errors for the coefficients by bootstrapping, treating each county across all dates as an observational unit for bootstrap resampling. We also built a dynamic version of the generalized linear model for cases and deaths as a proportion of the population to further investigate the effects of social distancing and other predictors that change daily. Final significance testing was based on bootstrapping to account for potential time-dependent correlation structures.

### Forecasting

For the accurate prediction of future COVID-19 cases and deaths, it is necessary to account for the fluid nature of the data streams comprising the PVI. Accordingly, we developed a Bayesian spatiotemporal random-effects model that jointly describes the log-observed and log-death counts to build local forecasts. Log-observed cases for a given day are predicted using known covariates (e.g., population density, social distancing metrics), a spatiotemporal random-effect smoothing component, and the time-weighted average number of cases for these counts. This smoothed time-weighted average is related to a Euler approximation of a differential equation; it provides modeling flexibility while approximating potential mechanistic models of disease spread. The smoothed case estimates are used in a similar spatiotemporal model that predicts future log-death counts based on a geometric mean estimate of the estimated number of observed cases for the previous seven days as well as the other data streams. The Dashboard shows the resulting county-level predictions and corresponding confidence intervals (Fig. 1). Details of the model are provided in the Supplemental Information.

**Figure 3.**
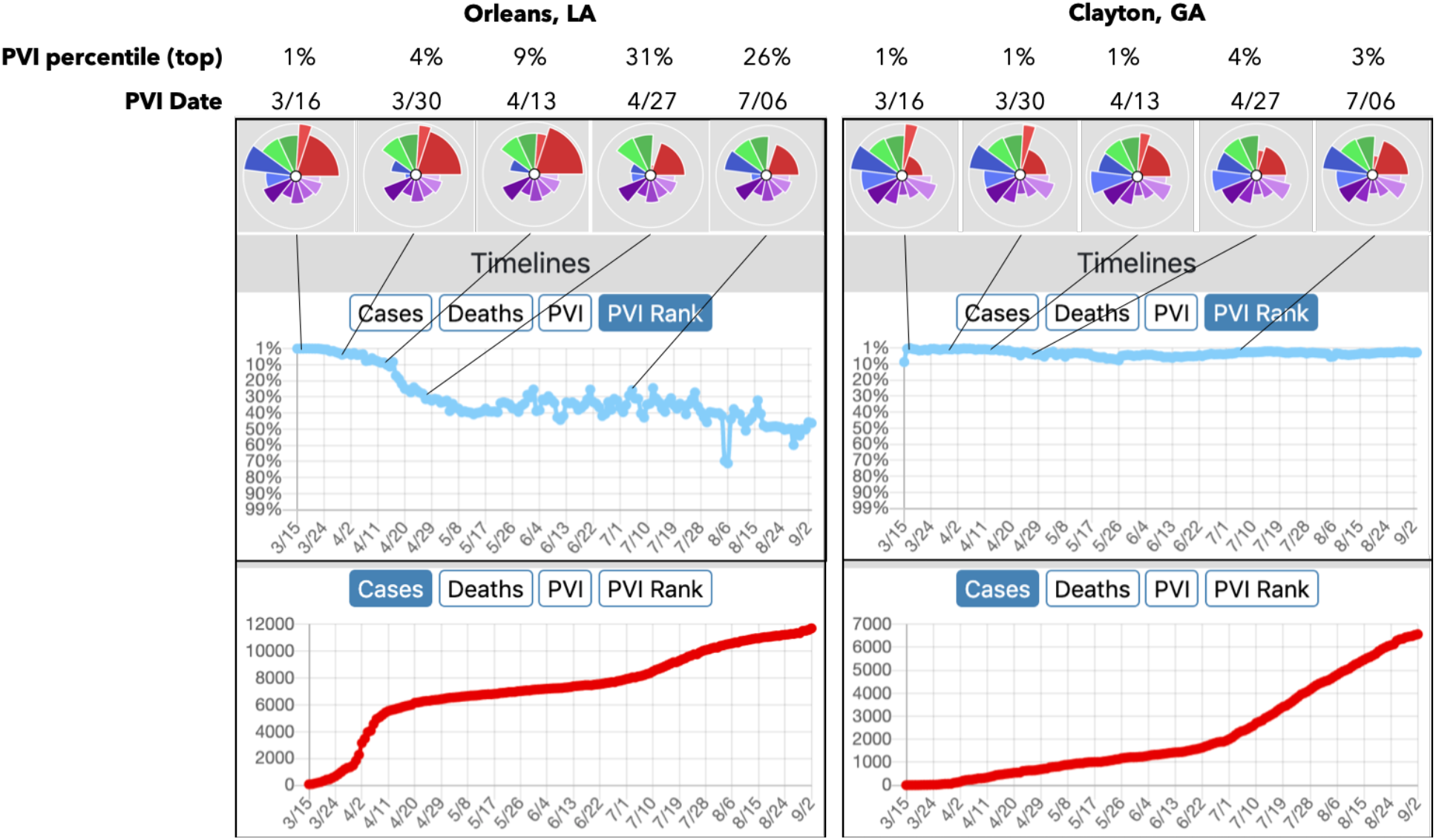
PVI data from the PVI Dashboard are shown for Orleans County, LA (left) and Clayton County, GA (right). The PVI profiles from March 15^th^, 2020 are shown above the timelines for each county. Comparing Orleans Parish (County) to Clayton County, they have similar ranks for Population Concentration (green slices) and Health and Environment (purple slices). For Intervention Practices (blue slices), differences can be seen; Orleans County’s adherence to social distancing measures and increased COVID-19 testing (reverse-scored and indicated by smaller blue slices) is visualized compared to Clayton. The changes in overall PVI rank across the timeline of the pandemic are shown. While the trajectory of new cases was blunted in Orleans County, it continued increasing in Clayton. Note that both counties observed spikes in the Cases trajectory as Social Distancing measures were relaxed at the end of June.

### Dashboard Technical Details

We used the ArcGIS JavaScript API (v4.13) (ESRI 2020) and custom PHP and JavaScript files to build the Dashboard web application. The API is used to overlay county borders, COVID-19 count data, and PVI model images on a Basemap or custom WebMap. County boundary data is from a feature service, while all other data is stored in an SQL database. PVI model images are base64-encoded scalable vector graphics (SVGs) rendered as inline images. The custom scripts controlling the Dashboard’s functionality optimize the efficiency of HTTP requests and other computational overhead to promote real-time interactivity. The Dashboard is hosted by the National Institute of Environmental Health (NIEHS) Office of Scientific Computing, which provides high-availability HTTPS load balanced with NGINX and a secure environment for web applications. Automated data updates are pushed to the public servers daily, and the daily update process is paralleled on a private server to permit independent data integrity assessment.

Complete, continually-updated documentation is available from a link on the Dashboard to a Quick Start Guide that introduces the PVI and Dashboard tools (https://www.niehs.nih.gov/research/programs/coronavirus/covid19pvi). A Details page provides additional in-depth information (https://www.niehs.nih.gov/research/programs/coronavirus/covid19pvi/details/index.cfm). The Supplemental Information and Figure S2 explain in detail the main features. The Dashboard is also mirrored as an iFrame on the CDC’s COVID Data Tracker, under the Unique Populations tab (https://covid.cdc.gov/covid-data-tracker/).

## Results

### PVI and Vulnerability

The summarization and communication goals of the PVI and the corresponding scorecards are human-centric and designed to convey and distill high-dimensional complex data. Our PVI model communicates an integrated concept of vulnerability that accounts for both dynamic (infection rate and interventions) and static (community population and healthcare characteristics) drivers. To gauge the association of daily PVI scores versus observed death outcomes, we assessed the rank-correlation between the overall PVI and the key vulnerability-related outcome metrics of cumulative deaths (Figure S1A), population adjusted cumulative deaths (Figure S1B), and the proportion of deaths from cases (Figure S1C). The Spearman Rho values for the PVI (from March 15 to August 12th) versus outcomes 1, 7, 14, 21, and 28 days ahead of a given day are displayed. All daily rank-correlation estimates were highly significant (p-values < 5.1E-14). The mean Rho values increase with a longer time horizon (blue text on Supplementary Figures 1A, 1B, 1C) and thus a highly-ranked PVI provides evidence that local actions should be taken to mitigate undesirable outcomes.

### Epidemiological Modeling

Supplementary Tables 2-7 display the regression coefficients produced with generalized linear modeling in a cross-sectional analysis of county cases and deaths up to 8/24/2020. As expected, the most significant predictor for the case count is population size (p<1E-300). The next most significant predictors associated with case counts are the proportion of black residents (p=1.28E-61) and mean PM_2.5_ (p=9.08E-32), followed by Insurance percent coverage (positively associated, p=1.51E-27) and proportion of Hispanic residents (p=6.92E-20) (Supplementary Table 2). In addition, the proportion of the population tested for SARS-Cov-2 infection is associated with case counts (p=3.39E-13), which we attribute to statewide responses to emerging infection clusters. In this cumulative analysis, social distancing and travel-related predictors were significant even though they represent aggregate values per county. For deaths as a proportion of the county population (Supplementary Table 4), the same predictors are highly significant, although the Insurance percent coverage is much less significant than for cases (p=4.48E-05). We note that cases and deaths per county population represent overall societal risk, for which vulnerability measures are relevant. The rank correlation coefficients for the PVI vs. the fitted values for the number of cases and the death rate are 0.54 and 0.55, respectively (*p*<10^-16^ for both).

Our analysis of the proportion of deaths per cases is enlightening, despite the previously noted caveat regarding potential bias due to testing variation. We note that a true case fatality rate potentially involves very different predictors than a case rate model, and deaths per county population are also closely tied to case rates. Our modeling (Supplementary Table 5) shows that after multiple test corrections, state testing rates are no longer significant in comparison to multiple-testing thresholds (p=0.036), which is consistent with the hypothesis that testing uncovers cases but does not predict case fatalities. The proportion of black residents (p=2.64E-12) and mean PM_2.5_ (p=1.92E-04) were significant, but less so than in the deaths/population model. Deaths/reported cases were found to be associated with the proportion of owner-occupied residences (p=9.31E-07) and inversely associated with median house value (p=0.000863). Both measures tend to be associated with wealth, but the relationship is complicated by the fact that high housing prices impede home ownership.

The results of the dynamic model for cases (Supplementary Table 6) with bootstrapped p-values had much stronger significance than the results of the cumulative case model, which we attribute to the dynamic model’s ability to account for additional sources of variation due to the use of lagged case counts, a smooth time-dependent term to account for national trend, and the inclusion of daily dynamic predictors. Again, the most significant predictors are the population size (*p*<1E-300), the proportion of black residents (*p*<1E-300), the two-week-lagged cumulative number of cases as a predictor of current cases (*p*<1E-300), and mean PM_2.5_ (*p*<1E-300). We also ran the analogous model for deaths/population size (Supplementary Table 7) and the same predictors were found to be highly significant. In summary, the dynamic versions of the generalized linear model reinforce and amplify the conclusions from the previous cumulative models. However, the models are not designed to perform forecasting, which can be viewed as essentially a machine learning exercise. For forecasting, careful cross-validation approaches can be used to assess the accuracy of the results.

The most consistent significant predictors for COVID-19 related case rates and mortality are the proportion of black residents and the mean PM_2.5_, reinforcing conclusions from previous reports (Dong, Du, & Gardner 2020). A one-percentage-point increase in the proportion of black residents is associated with a 2.9% increase in the COVID-19 death rate. The effect of a 1 g/m^3^ increase in PM_2.5_ is associated with an approximately 14.5% increase in the COVID-19 death rate, which is at the high end of a previously reported confidence interval from a report in late April 2020 (Wu et al. 2020) when deaths had reached 38% of the total as of June 2020. We find that these effects persist when including numerous additional predictors and correcting for factors such as socioeconomic status, housing density, and comorbidities. Moreover, the effects persist for a range of response values, including cumulative (i) cases, (ii) deaths, (iii) deaths as a proportion of the population, and (iv) deaths as a proportion of reported cases (Supplemental Tables 2-5). These results strongly suggest the important role of structural variation by location, which results in drastic health disparities. The results of the dynamic version of the generalized linear model (Supplemental Tables 6-7) support the importance of social vulnerability indicators and may be viewed as a sensitivity test that the impact of social distancing and other dynamic measures do not alter the significance of many of the social vulnerability indicators.

### Functional Power-Adjusted Relative Rate Model Forecasting

Data-driven machine learning was implemented for near term predictions of case and death outcomes at a local level. The resulting county-level predictions and corresponding confidence intervals for the next seven days are shown in the Dashboard ‘Predictions’ element. Additional details of the model and implementation are included in the Supplemental Information. The accuracy of the predictions were assessed by calculating the Pearson Correlation (Rho) of the predicted values versus the observed from June 3 through August 5, 2020. To avoid weekend-related reporting variation in cases and deaths, both predictions and observed cases/deaths were summed to weeks based on Wednesday through Tuesday. The accuracy of county-level predictions of Covid19-cases and -deaths were assessed by calculating the Pearson Correlation (Rho) of weekly predicted values versus weekly actual values across U.S. counties. For the 10 forecasts of Covid19 cases made each Wednesday from June 3rd through August 5th, the median Rho for 1-week out was 0.96. For deaths, the 1-week out median Rho for these 10 periods was 0.88. Summary Rho distributions are shown in Supplemental Figure 3, and scatterplots for all counties for the most recent analysis week are shown in Supplemental Figure 4.

### Dashboard Features

The interactive visualization within the PVI Dashboard communicates factors underlying vulnerability and empowers community action. On loading, the Dashboard displays the top 250 PVI profiles (by rank) for the current day. The data, PVI scores, and predictions are updated daily, and users can scroll through historical PVI and county outcome data. Individual profiles are an interactive map layer with numerous display options and filters that allow sorting by overall score and combinations of slice scores, clustering by profile similarity (i.e., vulnerability “shape”), and searching for counties by name or state. Any user-selected county overlays the summary Scorecard and populates the surrounding panels with county-specific information (Figure 1). Scrollable panels on the left include plots of vulnerability drivers relative to their nationwide distribution across all U.S. counties, with the location of the selected county delineated. The panels across the bottom of the Dashboard report cumulative county numbers of cases and deaths; timelines of cumulative cases, deaths, PVI scores, and PVI ranks; daily changes in cases and deaths for the most recent 14-day period (a measure commonly used in reopening guidelines); and predicted cases and deaths for a seven-day forecast horizon.

Taken together, the Dashboard features support the interactive evaluation and visualization of current data for localities while providing context with respect to all U.S. counties. Full time series of case, death, and PVI trends enable the examination of the track records of counties of interest as well as the comparison of trajectories for peer, or comparable, counties in terms of varying success with specific interventions. For example, using Orleans County (home to New Orleans, LA) as an exemplar, we employed the multi-criteria filtering capabilities in the Dashboard to find a peer county for comparison. By bounding the PVI to similar ranges of vulnerability drivers (i.e., slices) for population mobility, residential density, and population demographics, we identified a subset of candidate counties and ultimately chose Clayton County, GA to illustrate the effects of dramatic differences in public action/interventions. Figure 3 shows detailed results for the two counties, which have similar baseline vulnerabilities but implemented divergent interventions at the outset of the pandemic. Specifically, pronounced differences in intervention measures (social distancing and testing) are associated with varying dynamics of the infection rates in these counties, as visualized through the considerable differences in magnitude of the blue (intervention-related) and red (infection-related) slices over time. Note that all data streams are scaled so that a larger slice indicates increased vulnerability (e.g., the larger blue slices represent less adherence to social distancing and lower testing rates). As visualized in Figure 3, the PVI rank for Orleans County improves over time (i.e., follows a downward rank/percentile path), effectively blunting the curve caused by the accelerated increase in the number of cases through early interventions. There is no similar positive change for Clayton County, reflecting differences due to varying interventions in the two areas. In this way, the PVI Dashboard enables customized empirical comparisons and evaluations across peer counties.

## Discussion

Numerous expert groups have coalesced around a general roadmap to address the current COVID-19 pandemic that comprises (i) reducing the spread through social distancing, (ii) gradually easing restrictions while monitoring for resurgence and healthcare overcapacity, and (iii) eventually moving to pharmaceutical interventions. However, the responsibility for navigating the COVID-19 response falls largely on state and local officials, who require data at the community level to make equitable decisions on allocating resources, caring for vulnerable sub-populations, and enhancing/relaxing social distancing measures. The goal of the COVID-19 PVI Dashboard is to empower informed actions to combat the pandemic from the local to the national levels on multiple time scales. The Dashboard accomplishes this goal by combining underlying COVID-19-specific structural vulnerabilities with dynamic infection and intervention data at the county level to produce an integrated concept of vulnerability that can inform decision-making on actions at the local level.

Furthermore, the general public must embrace interventions for them to be effective, and interactive visualization is a proven approach to communication among diverse audiences. The PVI Dashboard provides interactive, visual profiles of vulnerability atop an underlying statistical framework that enables the comparison of counties by clustering and the evaluation of the PVI’s sensitivity to component data. The Dashboard’s county-level Scorecards illustrate both overall vulnerability and the components driving it. A key utility of a public-facing, interactive dashboard is that decision-makers can point to it for support, thus promoting transparency and public buy-in for actions taken in the interest of public health. Example use cases include the priority distribution of medical resources such as hospital beds, targeted community outreach activities, and the establishment of contact-tracing mechanisms. Eventually, the PVI could be used to support the priority distribution of vaccines to highly-vulnerable communities.

The modeling efforts presented here support decision-making in multiple ways. The epidemiological modeling enables testing the impact of changes in dynamic interventions, such as changes in social distancing, and the forecasting efforts support short-term resource allocation decisions, such as hospital staffing and the distribution of supplies. These forecasts also help communicate the trends that are part of the CDC’s reopening criteria (Centers for Disease Control and Prevention 2020), such as whether interventions and local government actions translate into flattened curves. The PVI score itself constitutes an integrated indicator of vulnerability that is strongly associated with mortality outcomes in the near-to-medium term.

The overall PVI score highlights highly-ranked counties that should consider taking local actions or receive targeted help to mitigate undesirable outcome trends. The *Timelines* panel of the Dashboard illustrates observed county-level changes over time to answer questions such as “Have we flattened the curve?” while the *Predictions* panel presents statistically robust forecasts that consider all of the data to answer the question, “What’s next?”. Overall, the COVID-19 PVI Dashboard can help facilitate decision-making and communication among government officials, scientists, community leaders, and the public to enable more effective and coordinated action to combat the pandemic.

A growing number of risk factors have been highlighted in the rapidly expanding scientific studies on COVID-19. Infectious disease epidemiology is rapidly evolving, and new risk factors and environmental variables (e.g., weather conditions) are continually being discovered. From the early identification of older individuals’ vulnerability (Verity et al. 2020) to the dramatic racial disparities that have been more recently highlighted (Coughlin et al. 2020), there is mounting evidence that socially marginalized populations are suffering disproportionally. While this is not unique to the ongoing pandemic, it is clear that pandemic vulnerability and response are dynamic and differ across communities (Quinn & Kumar 2020). The COVID-19 PVI Dashboard provides contextualized local summaries of differences in vulnerability and highlights racial disparities, even when adjusting for multiple covariates. The Dashboard’s presentation of information in a relative sense enables the fair comparison of communities of different sizes to support prioritization decisions. Further, the PVI visualization is a human-centric and communicates how particular communities’ drivers (i.e., slices) differ in terms of their contribution to overall vulnerability. This visualization promotes transparency by clarifying the judgments and trade-offs entering into such an assessment while maintaining a direct link to the underlying quantitative data.

COVID-19 will continue to present public health challenges into the foreseeable future. By integrating vulnerability information (both historical and forward-looking), the Dashboard supports key decision-making for managing the ongoing pandemic. We will continue to update the data streams combined to calculate the PVI and will add additional variables as evidence of new risk factors and potential drivers of vulnerability emerge and supported by publicly available data. We will also continually develop software tools so that people can actively build their own models and will update the modeling efforts as needed. Combating endemic diseases requires long-range thinking, informed action, and political will, and we offer the COVID-19 PVI Dashboard as an interactive monitoring tool to support these sustained efforts.

## Data Availability

All data is publicly available at https://github.com/COVID19PVI/data

https://covid19pvi.niehs.nih.gov/

## Acknowledgments

We would like to thank the IT and web services staff at NIEHS for their help and support, as well as JK Cetina and DJ Reif for useful technical input and advice.

## Author contributions

*Concept and design:* SM, FW, WC, IR, AMR, DR. *Acquisition, analysis, or interpretation of data:* All authors. *Drafting of the manuscript:* All authors. *Critical revision of the manuscript for important intellectual content:* All authors. *Statistical analysis:* JH, MW, KS, YZ, AMR, DR. *Obtained funding:* WC, IR, DR, AMR.

## Data and materials availability

All data is available through links in the main text or the supplementary materials.

